# Prevalence of chronic kidney disease in the general population in Latin America and the Caribbean: Protocol for a systematic review and meta-analysis

**DOI:** 10.1101/2021.01.26.21250540

**Authors:** Diego A Sequeiros-Buendia, Camila S Villa-Ato, Marlies Weiss-Carlini, Rodrigo M Carrillo-Larco

## Abstract

**Background:** Chronic kidney disease (CKD) is a global health issue with a general prevalence of 9%. Although the most affected populations are in low- and middle-income countries, the epidemiology of CKD in these countries remains poorly understood and prevalence estimates come from global efforts informed by data from high-income countries; these prevalence estimates need to be compared –and if needed updated–with local estimates.

**Objective:** To estimate the prevalence of CKD in adults in Latin America and the Caribbean (LAC).

**Methods:** Systematic review and meta-analysis. We will search Embase, Medline, Global Health (these three through Ovid), Scopus and LILACS. No date or language restrictions will be set. We seek observational studies with a random sample of the general population. We will screen titles and abstracts, we will then study the selected reports. Both phases will be done by two reviewers independently. Data extraction will be performed by two researchers independently using a pre-specified Excel form. We will evaluate the risk of bias with the scale proposed by Hoy et al. for prevalence studies. We will conduct a meta-analysis of prevalence estimates, if there are at least three reports homogeneous enough to be pooled; we will use a random-effects model.

**Conclusions:** This systematic review and meta-analysis will provide the prevalence of CKD in adults in countries of LAC. Currently, information regarding CKD in the region is limited. This work will provide evidence to elucidate the magnitude of CKD prevalence in LAC. In so doing, we will provide evidence to inform the scientific community about the burden of CKD in LAC so that research, policies and health interventions can be planned accordingly.

## BACKGROUND

Chronic kidney disease (CKD) is a global health issue disproportionately affecting low- and middle-income countries like those in Latin America and the Caribbean (LAC).(1– 3) The overall CKD prevalence in LAC has been estimated at 10%, slightly above the global prevalence (9%).(4) Similarly, though with much higher magnitude, in LAC CKD mortality and disability metrics are higher than the global estimates.(5) Although these figures teach about the burden of CKD in LAC, they are mostly informed by data from high-income countries where risk factors levels and access to health-care may be different,(6–11) thus prevalence estimates would not be equivalent. Whether population-based (national) studies in LAC agree with these international metrics, is largely unknown.(12) A systematic and comprehensive search of all relevant data in LAC could complement and further inform international efforts so that they better estimate the epidemiology of CKD in LAC.

A thorough understanding of the CKD epidemiology in LAC is much needed to inform research priorities and data needs (e.g., where research/data are needed), to inform clinical guidelines with basic epidemiological metrics (e..g, prevalence), and to provide insights for policies and interventions (e.g., whom and where would benefit the most). Nonetheless, to begin an exhaustive characterization of the CKD epidemiology in LAC, the prevalence needs to be well estimated, along with determinants such a sex, age and geographic variations. There have not been any LAC-wide studies to provide this information, and individual efforts, either at the national and community levels, have not been systematically gathered, appraised and if possible pooled to provide a metric for LAC as a region.

We aim to quantify and appraise the prevalence of CKD in LAC, through a systematic review and meta-analysis of population-based studies conducted in LAC. We will deliver a comprehensive appraisal of the available evidence, signalling research gaps and pinpointing research needs; also, if methodologically appropriate, we will deliver pooled prevalence estimates of CKD for LAC and by sub-regions.

## METHODS

### Objective

To estimate the prevalence of CKD in the general population of LAC.

### Study design

This systematic review and meta-analysis will be conducted following the preferred reporting items for systematic reviews and meta-analyses (PRISMA) guidelines.(13)

### Eligibility criteria

#### Participants/population

We will include men and women of the general population aged 18 years and above. Participants should have been selected following any kind of random sampling technique. We will only include people in countries in LAC. Overall, we aim to include original reports in which the study population resembles -as much as possible-the general population.

#### Intervention, exposure

No intervention will be studied. No specific exposure will be included.

#### Comparator/control

Not applicable.

#### Outcome

Prevalence of CKD.

#### Types of studies

We will include population-based epidemiological observational studies: cross-sectional studies, (national) surveys, and cohort studies (baseline). We will include reports in which a random sample of the general population in LAC was studied to assess the prevalence of CKD. We will include studies in which CKD was defined as the presence of kidney damage indicated by urine albumin-creatinine ratio, urine protein-creatinine ratio, albumin excretion ratio, urine sediment, kidney images, kidney biopsy or any combination of these, or alteration of the glomerular filtration rate (GFR) estimated by using serum creatinine and/or serum cystatin C, regardless of the formula used to compute the estimated glomerular filtration rate (eGFR). We will select original research reports including research letters and abstracts, as long as they provide enough information to evaluate the selection criteria.

### Exclusion criteria

We will exclude the following study designs: case-control, case reports, editorials, commentaries, narrative and scoping reviews, clinical trials, any grey literature (e.g., dissertations/thesis), and systematic reviews/meta analyses. If we retrieve any systematic review on this subject, we will revise its reference list to identify relevant original sources. We will exclude studies with LAC populations outside the LAC region (e.g., Latins in the USA). We will exclude studies which only sampled people under 18 years of age, those in which the outcome was ascertained based on self-reported history of CKD only, and studies which focused on individuals with a specific risk factor (e.g., consumers of nephrotoxic drugs) or condition (e.g., hypertension).

If there are any original reports in which the population, methodology or results are not clear enough to assess our inclusion/exclusion criteria, we will contact the corresponding author by email. We will wait a reasonable time (two weeks), if we receive no answer and cannot solve our doubts through other means, this report will be excluded based on the lack of clarity to assess inclusion/exclusion criteria.

### Literature search and data collection

– The search will be conducted in five search engines: Embase, Medline, Global Health (these three through Ovid), Scopus and LILACS. No date or language restrictions will be set. The complete search strategy can be found in Appendix I.
– Titles and abstracts will be screened by two researchers independently, looking for studies that meet the selection criteria above detailed. After this initial phase, full-text reports of the selected publications will be studied by two researchers independently. Any discrepancies at any stage will be solved by consensus or by a third party.
– We will record the reasons for exclusion in the full-text phase and summarize the number of included/excluded reports following the PRISMA flow diagram.

### Data extraction

We will develop a data extraction form in an Excel file. We will pilot this form with a random sample of ten selected publications. After this pilot phase, we will update the extraction form as needed; the extraction form will not be modified thereafter. Data extraction will be conducted by two researchers independently; discrepancies will be solved by consensus or by a third party. We anticipate to extract the following information:

– Reference: First author, article title, country where the study took place, year of publication, year of data collection and journal name.
– Study population: Number of participants included in the results, characteristics of the study population including proportion of female subjects and mean age, original inclusion/exclusion criteria, and sampling technique.
– Outcome: Method to ascertain CKD diagnosis, overall CKD prevalence along with uncertainty estimate (e.g., 95% confidence interval). If available, we will also extract the prevalence estimate by gender. Similarly, we will extract mean eGFR and if available by gender.

If we found multiple reports with the same underlying data (e.g., an open-access national survey analysed by multiple researchers), we will include only one report. That is, we will select the report with the largest sample size or the one providing most information (e.g., overall prevalence along with sex-stratified estimates).

### Risk of bias and Methodological Quality of Studies

We will use the risk of bias tool developed by Hoy and colleagues for prevalence studies.(14) The items of this tool will be implemented in a spreadsheet, and two reviewers, independently, will complete the information for each selected report. Discrepancies will be solved by consensus or a third party. This process will not influence the planned qualitative and quantitative synthesis.

### Statistical analysis

We plan to report our findings in two ways: qualitative and quantitative. First, we will summarise the main items extracted from each report in tables and figures as relevant. We will use this information to narratively describe our findings and the characteristics of the selected reports. Second, we plan to conduct a random effects meta-analysis, if there are at least three (≥3) original reports with sufficient information and homogenous enough to pool. In this way, we will provide a pooled prevalence estimate of CKD in LAC; if possible, we will stratified this pooled prevalence estimate by sex and sub-region (e.g., the Caribbean vs South America). We will use R or STATA statistical software to conduct the random effects meta-analysis with the DerSimonian and Laird method. We will assess publication bias through funnel plots and the Egger’s test; we propose a minimum of ten original reports to study publication bias. A p<0.05 will be considered statistically significant throughout the analysis plan.

## Data Availability

None to declare

## APPENDIX

**Appendix 1:**
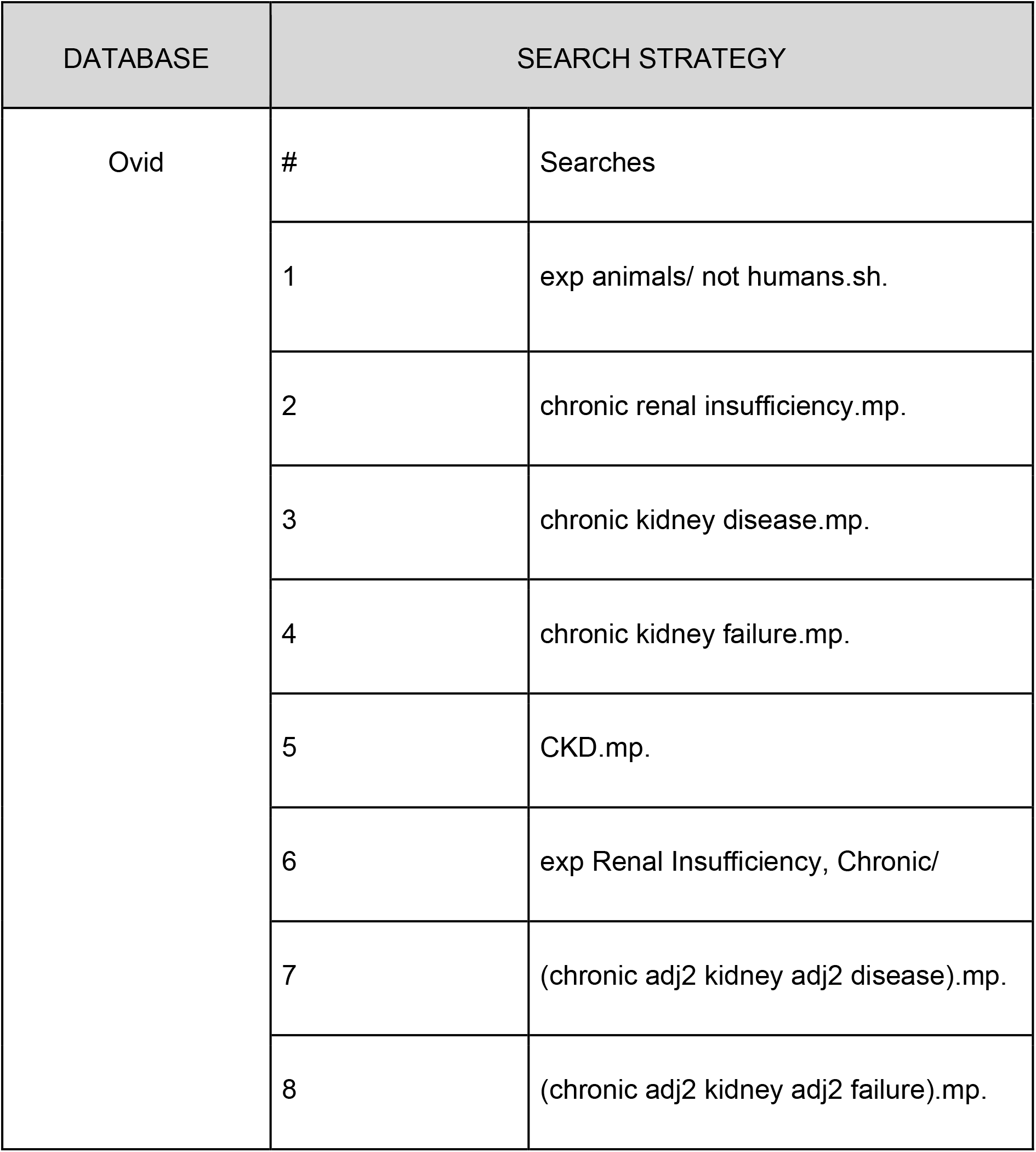
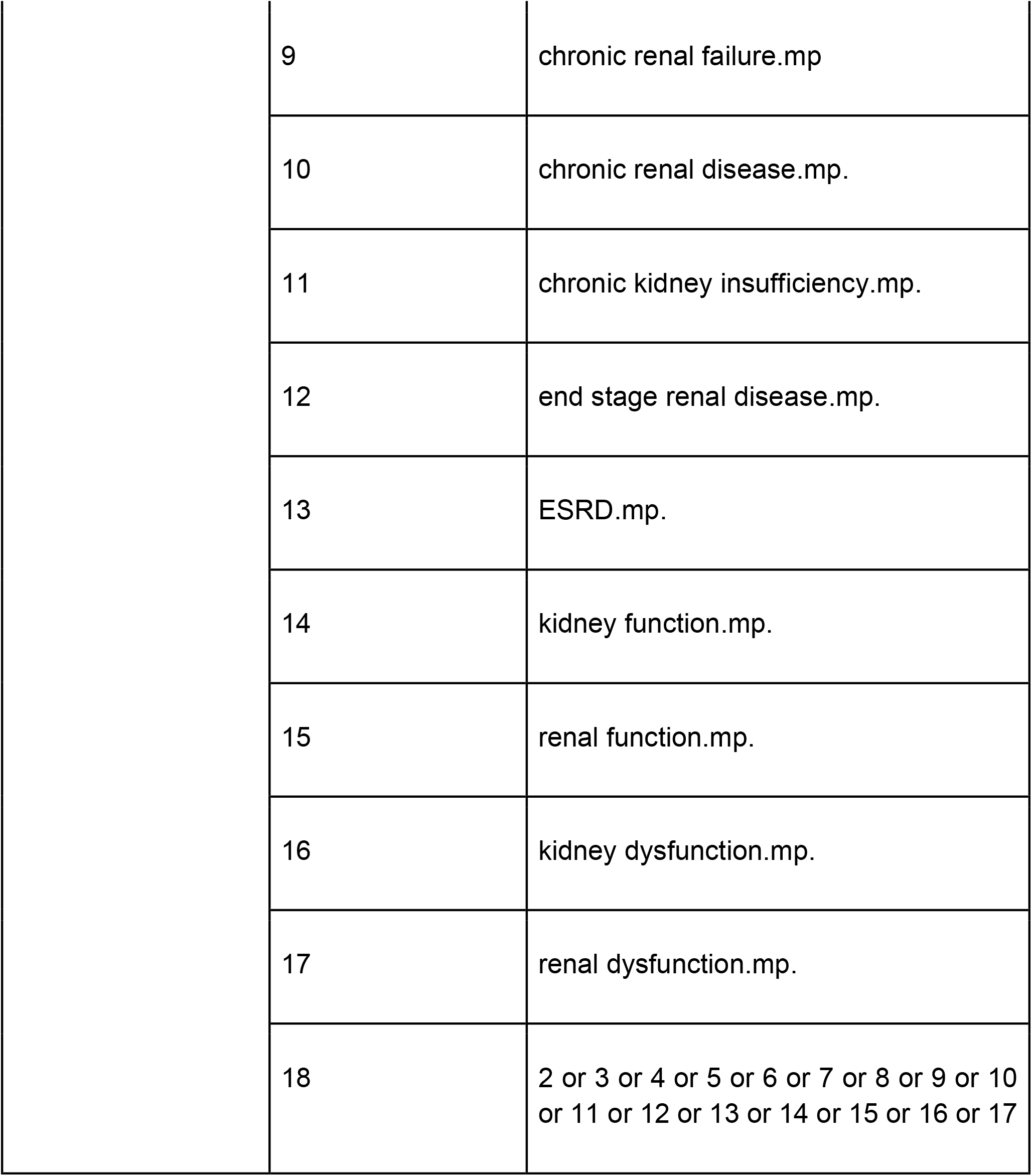
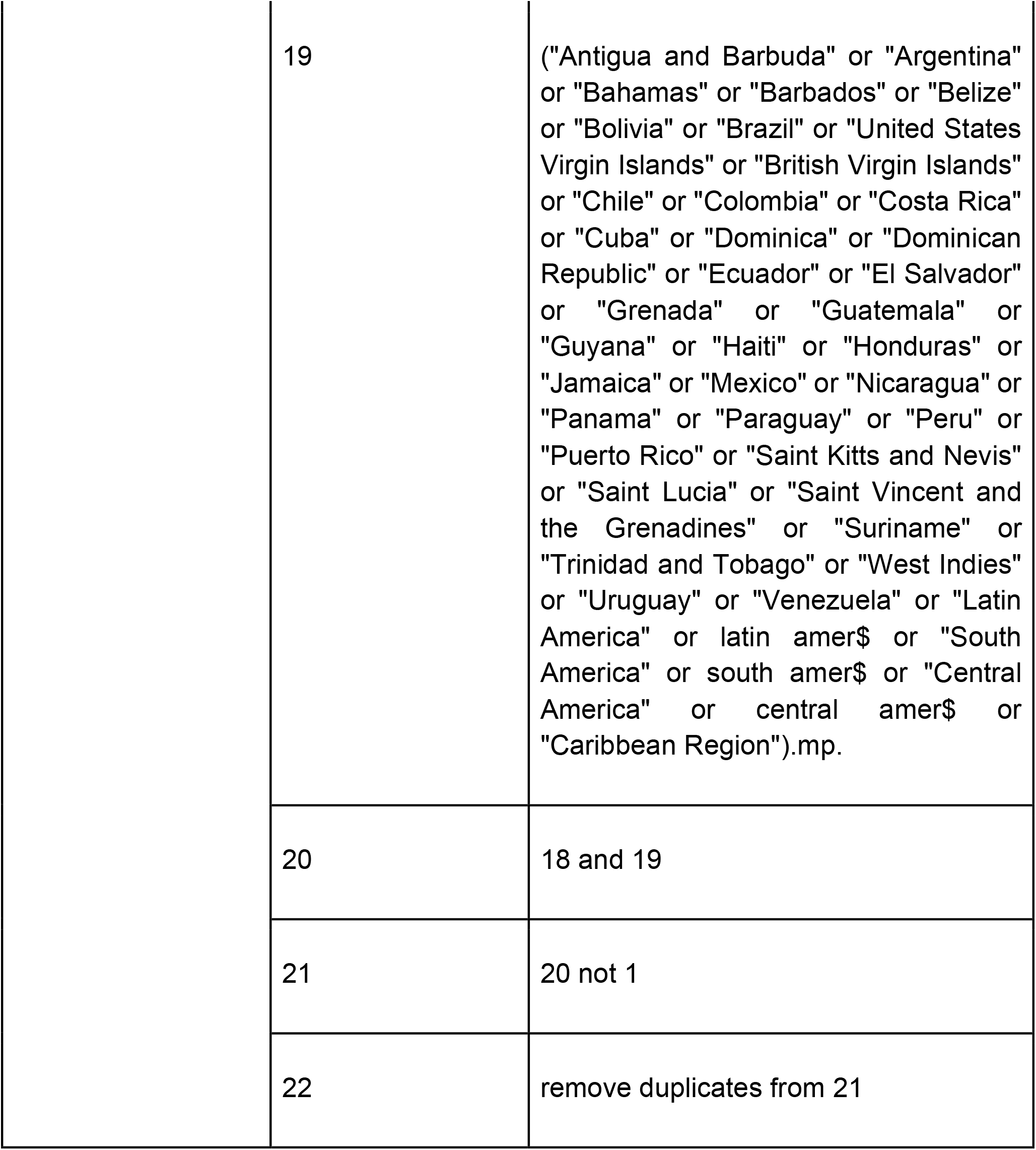
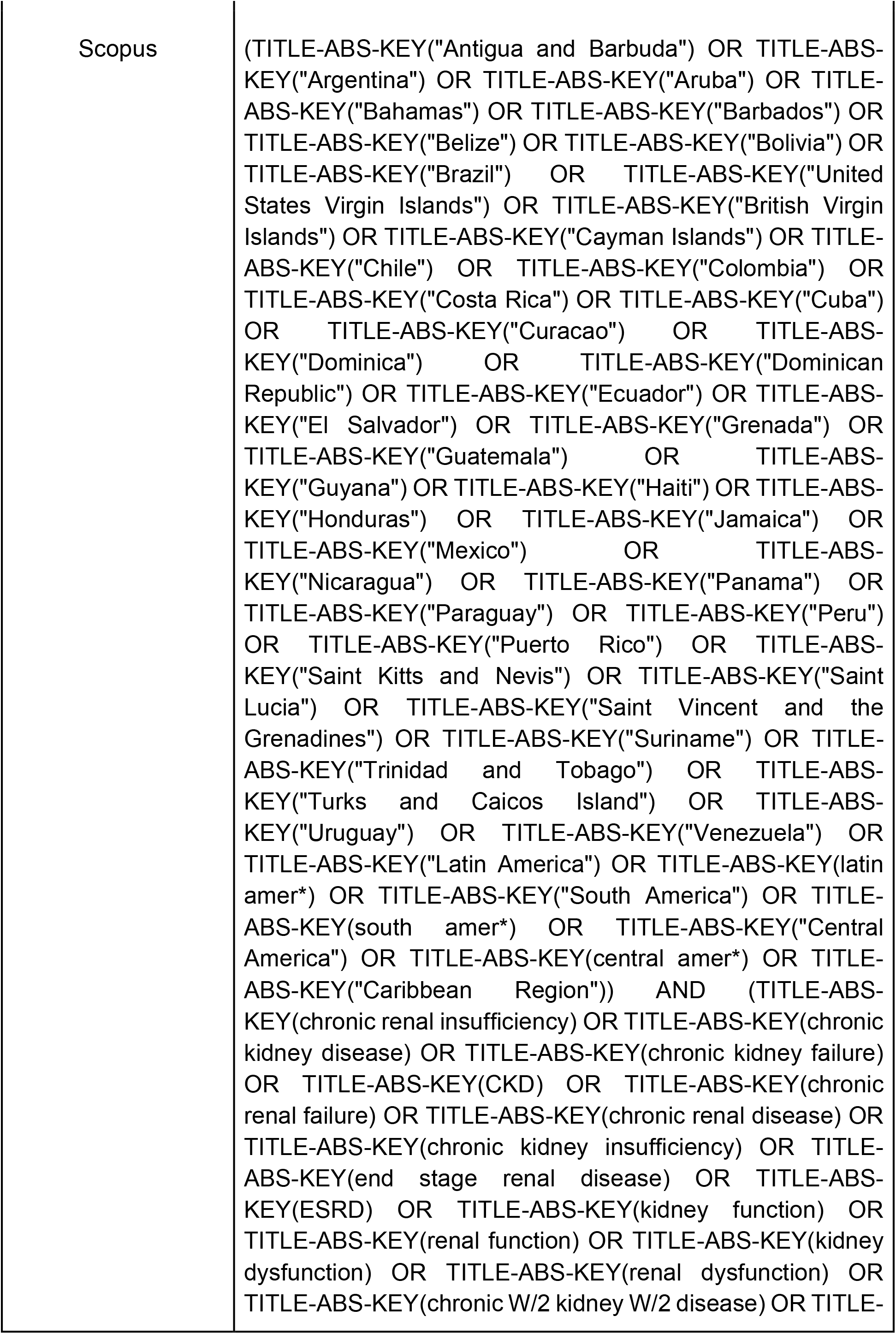
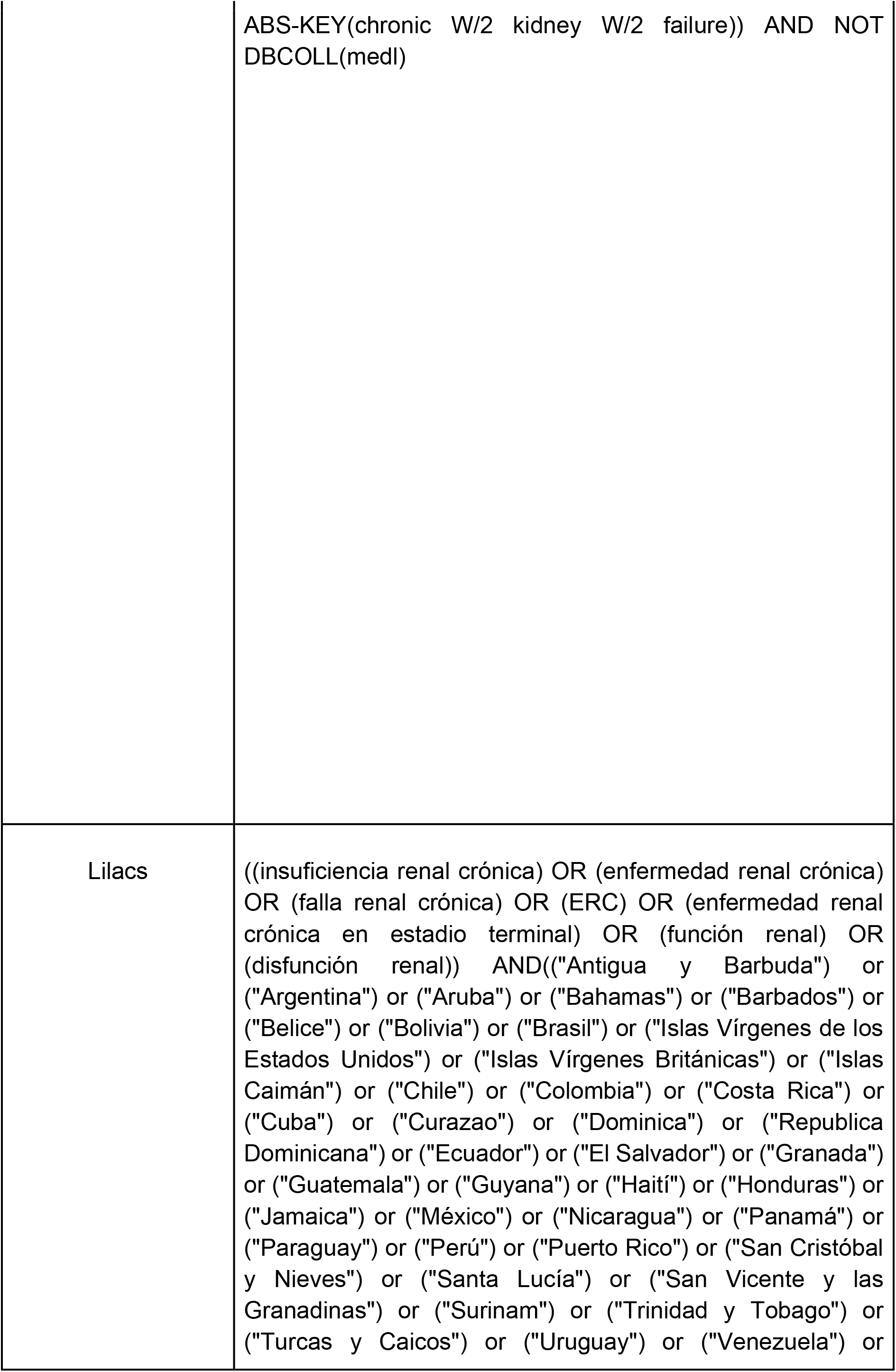
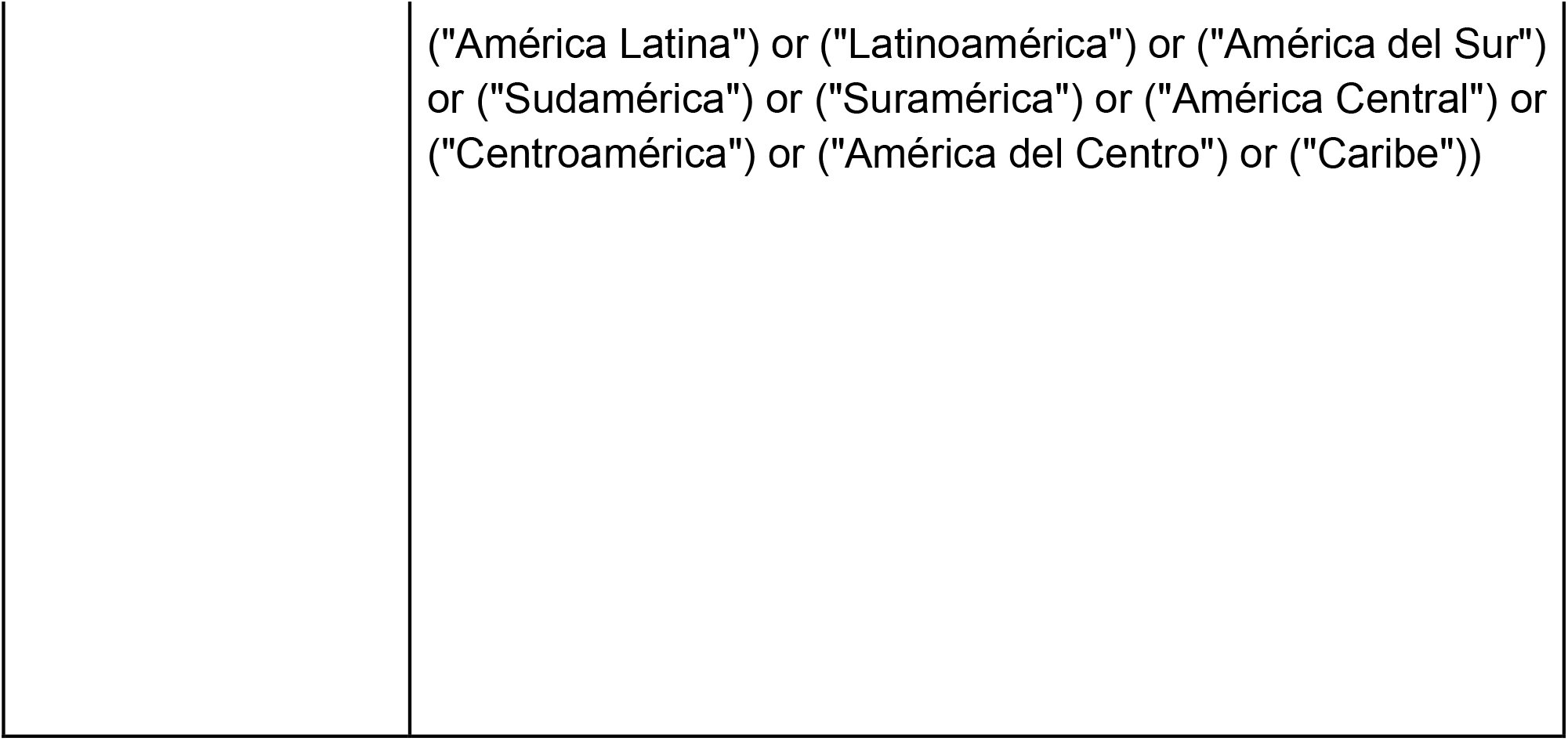
Search strategy.

| DATABASE | SEARCH STRATEGY |  |
| --- | --- | --- |
| Ovid | # | Searches |
|  | 1 | exp animals/ not humans.sh. |
|  | 2 | chronic renal insufficiency.mp. |
|  | 3 | chronic kidney disease.mp. |
|  | 4 | chronic kidney failure.mp. |
|  | 5 | CKD.mp. |
|  | 6 | exp Renal Insufficiency, Chronic/ |
|  | 7 | (chronic adj2 kidney adj2 disease).mp. |
|  | 8 | (chronic adj2 kidney adj2 failure).mp. |

|  |  |
| --- | --- |
| 9 | chronic renal failure.mp |
| 10 | chronic renal disease.mp. |
| 11 | chronic kidney insufficiency.mp. |
| 12 | end stage renal disease.mp. |
| 13 | ESRD.mp. |
| 14 | kidney function.mp. |
| 15 | renal function.mp. |
| 16 | kidney dysfunction.mp. |
| 17 | renal dysfunction.mp. |
| 18 | 2 or 3 or 4 or 5 or 6 or 7 or 8 or 9 or 10<br>or 11 or 12 or 13 or 14 or 15 or 16 or 17 |
| | 19 | ("Antigua and Barbuda" or "Argentina" or "Bahamas" or "Barbados" or "Belize" or "Bolivia" or "Brazil" or "United States Virgin Islands" or "British Virgin Islands" or "Chile" or "Colombia" or "Costa Rica" or "Cuba" or "Dominica" or "Dominican Republic" or "Ecuador" or "El Salvador" or "Grenada" or "Guatemala" or "Guyana" or "Haiti" or "Honduras" or "Jamaica" or "Mexico" or "Nicaragua" or "Panama" or "Paraguay" or "Peru" or "Puerto Rico" or "Saint Kitts and Nevis" or "Saint Lucia" or "Saint Vincent and the Grenadines" or "Suriname" or "Trinidad and Tobago" or "West Indies" or "Uruguay" or "Venezuela" or "Latin America" or latin amer\$ or "South America" or south amer\$ or "Central America" or central amer\$ or "Caribbean Region").mp. |
|  | 20 | 18 and 19 |
|  | 21 | 20 not 1 |
|  | 22 | remove duplicates from 21 |

|  |  |
| --- | --- |
| Scopus | (TITLE-ABS-KEY("Antigua and Barbuda") OR TITLE-ABS-KEY("Argentina") OR TITLE-ABS-KEY("Aruba") OR TITLE-ABS-KEY("Bahamas") OR TITLE-ABS-KEY("Barbados") OR TITLE-ABS-KEY("Belize") OR TITLE-ABS-KEY("Bolivia") OR TITLE-ABS-KEY("Brazil") OR TITLE-ABS-KEY("United States Virgin Islands") OR TITLE-ABS-KEY("British Virgin Islands") OR TITLE-ABS-KEY("Cayman Islands") OR TITLE-ABS-KEY("Chile") OR TITLE-ABS-KEY("Colombia") OR TITLE-ABS-KEY("Costa Rica") OR TITLE-ABS-KEY("Cuba") OR TITLE-ABS-KEY("Curacao") OR TITLE-ABS-KEY("Dominica") OR TITLE-ABS-KEY("Dominican Republic") OR TITLE-ABS-KEY("Ecuador") OR TITLE-ABS-KEY("El Salvador") OR TITLE-ABS-KEY("Grenada") OR TITLE-ABS-KEY("Guatemala") OR TITLE-ABS-KEY("Guyana") OR TITLE-ABS-KEY("Haiti") OR TITLE-ABS-KEY("Honduras") OR TITLE-ABS-KEY("Jamaica") OR TITLE-ABS-KEY("Mexico") OR TITLE-ABS-KEY("Nicaragua") OR TITLE-ABS-KEY("Panama") OR TITLE-ABS-KEY("Paraguay") OR TITLE-ABS-KEY("Peru") OR TITLE-ABS-KEY("Puerto Rico") OR TITLE-ABS-KEY("Saint Kitts and Nevis") OR TITLE-ABS-KEY("Saint Lucia") OR TITLE-ABS-KEY("Saint Vincent and the Grenadines") OR TITLE-ABS-KEY("Suriname") OR TITLE-ABS-KEY("Trinidad and Tobago") OR TITLE-ABS-KEY("Turks and Caicos Island") OR TITLE-ABS-KEY("Uruguay") OR TITLE-ABS-KEY("Venezuela") OR TITLE-ABS-KEY("Latin America") OR TITLE-ABS-KEY(latin amer*) OR TITLE-ABS-KEY("South America") OR TITLE-ABS-KEY(south amer*) OR TITLE-ABS-KEY("Central America") OR TITLE-ABS-KEY(central amer*) OR TITLE-ABS-KEY("Caribbean Region")) AND (TITLE-ABS-KEY(chronic renal insufficiency) OR TITLE-ABS-KEY(chronic kidney disease) OR TITLE-ABS-KEY(chronic kidney failure) OR TITLE-ABS-KEY(CKD) OR TITLE-ABS-KEY(chronic renal failure) OR TITLE-ABS-KEY(chronic renal disease) OR TITLE-ABS-KEY(chronic kidney insufficiency) OR TITLE-ABS-KEY(end stage renal disease) OR TITLE-ABS-KEY(ESRD) OR TITLE-ABS-KEY(kidney function) OR TITLE-ABS-KEY(renal function) OR TITLE-ABS-KEY(kidney dysfunction) OR TITLE-ABS-KEY(renal dysfunction) OR TITLE-ABS-KEY(chronic W/2 kidney W/2 disease) OR TITLE- |
|  | ABS-KEY(chronic W/2 kidney W/2 failure)) AND NOT DBCOLL(medl) |
| Lilacs | ((insuficiencia renal crónica) OR (enfermedad renal crónica) OR (falla renal crónica) OR (ERC) OR (enfermedad renal crónica en estadio terminal) OR (función renal) OR (disfunción renal)) AND(("Antigua y Barbuda") or ("Argentina") or ("Aruba") or ("Bahamas") or ("Barbados") or ("Belice") or ("Bolivia") or ("Brasil") or ("Islas Vírgenes de los Estados Unidos") or ("Islas Vírgenes Británicas") or ("Islas Caimán") or ("Chile") or ("Colombia") or ("Costa Rica") or ("Cuba") or ("Curazao") or ("Dominica") or ("Republica Dominicana") or ("Ecuador") or ("El Salvador") or ("Granada") or ("Guatemala") or ("Guyana") or ("Haití") or ("Honduras") or ("Jamaica") or ("México") or ("Nicaragua") or ("Panamá") or ("Paraguay") or ("Perú") or ("Puerto Rico") or ("San Cristóbal y Nieves") or ("Santa Lucía") or ("San Vicente y las Granadinas") or ("Surinam") or ("Trinidad y Tobago") or ("Turcas y Caicos") or ("Uruguay") or ("Venezuela") or |
|  | ("América Latina") or ("Latinoamérica") or ("América del Sur") or ("Sudamérica") or ("Suramérica") or ("América Central") or ("Centroamérica") or ("América del Centro") or ("Caribe")) |

